# Predicting the future risk of lung cancer: development and validation of QCancer2 (10-year risk) lung model and evaluating the model performance of nine prediction models

**DOI:** 10.1101/2022.06.04.22275868

**Authors:** Weiqi Liao, Carol Coupland, Judith Burchardt, David Baldwin, the DART initiative, Fergus Gleeson, Julia Hippisley-Cox

**Affiliations:** Nuffield Department of Primary Care Health Sciences, University of Oxford, Oxford, UK; School of Medicine, University of Nottingham, Nottingham, UK; Nottingham University Hospitals NHS Trust, Nottingham, UK; Department of Oncology, University of Oxford, Oxford, UK

**Keywords:** lung cancer, risk prediction model, screening, early detection, diagnosis, low-dose computerised tomography (LDCT), targeted lung health check (TLHC) programme

## Abstract

**Objectives:** To develop and validate the QCancer2 (10-year risk) lung model for estimation of future risk of lung cancer and to compare the model performance against other prediction models for lung cancer screening

**Design:** open cohort study using linked electronic health records (EHRs) from the QResearch database (1 January 2005 – 31 March 2020)

**Setting:** English primary care

**Participants:** 12.99 million patients aged 25-84 years were in the derivation cohort to develop the models and 4.14 million patients were in the validation cohort. All patients were free of lung cancer at baseline.

**Main outcome measure:** Incident lung cancer cases

**Methods:** There were two stages in this study. First, Cox proportional hazards models were used in the derivation cohort to update the QCancer (10-year risk) lung model in men and women for a 10-year predictive horizon, including two new predictors (pneumonia and venous thromboembolism) and more recent data. Discrimination measures (Harrell’s C, D statistic, and 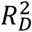) and calibration plots were used to evaluate model performance in the validation cohort by sex. Secondly, seven prediction models for lung cancer screening (LLP_v2_, LLP_v3_, LCRAT, PLCO_M2012_, PLCO_M2014_, Pittsburgh, and Bach) were selected to compare the model performance with the QCancer2 (10-year risk) lung model in two subgroups: (1) smokers and non-smokers aged 40-84 years and (2) ever-smokers aged 55-74 years.

**Results:** 73,380 incident lung cancer cases were identified in the derivation cohort and 22,838 in the validation cohort during follow-up. The updated models explained 65% of the variation in time to diagnosis of lung cancer 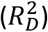 in both sexes. Harrell’s C statistics were close to 0.9 (indicating excellent discrimination), and the D statistics were around 2.8. Compared with the original models, the discrimination measures in the updated models improved slightly in both sexes. Compared with other prediction models, the QCancer2 (10-year risk) lung model had the best model performance in discrimination, calibration, and net benefit across three predictive horizons (5, 6, and 10 years) in the two subgroups.

**Conclusion:** Developed and validated using large-scale EHRs, the QCancer2 (10-year risk) lung model can estimate the risk of an individual patient aged 25-84 years for up to 10 years. It has the best model performance among other prediction models. It has potential utility for risk stratification of the English primary care population and selection of eligible people at high risk for the targeted lung health check programme or lung cancer screening.

**What is already known on this topic:**

- Using risk prediction models to stratify people at the population level and selecting those at the highest risks is an efficient and cost-effective strategy for screening programmes. It avoids waste of resources in screening patients at low risk.
- An ideal prediction model should have excellent discrimination and calibration in the target population.
- The Liverpool Lung Project (LLP_v2_) and the Prostate Lung Colorectal and Ovarian (PLCO_M2012_) models had only moderate discrimination and were not well-calibrated when externally validated using the Clinical Practice Research Datalink (CPRD) data for the English primary care population.

**What this study adds:**

- Developed and validated using robust statistical methodologies, the QCancer2 (10-year risk) lung model shows excellent discrimination and calibration in both sexes. It can estimate an individual adult patient’s risk for each year of follow-up, for up to 10 years.
- The QCancer2 (10-year risk) lung model has the best model performance in discrimination and calibration when compared with the other eight models (QCancer (10-year risk), LLP_v2_, LLP_v3_, LCRAT, PLCO_M2012_, PLCO_M2014_, Pittsburgh, and Bach) in three predictive horizons (5/6/10 years) and two sub-populations (smokers and non-smokers aged 40-84 years and ever-smokers aged 55-74 years).
- The QCancer2 (10-year risk) lung model can be applied to the English primary care population to select eligible patients for the Targeted Lung Health Check programme or lung cancer screening using low dose CT.

## Introduction

Lung cancer is a research priority in the UK. It is the third most common cancer in incidence and the most common cause of cancer death in the UK^1^. Lung cancer survival in the UK is worse than in other European countries (the EuroCare studies)^2^ and international counterparts (the ICBP studies)^3 4^. Research evidence from randomised controlled trials (RCTs) has shown that using low dose computerised tomography (LDCT) for lung cancer screening reduces mortality^5–7^. The United States Preventive Services Task Force (USPSTF) recommended using LDCT for lung cancer screening in 2013^8^, and relaxed the eligibility criteria for screening in 2021, by lowering the age threshold from 55 to 50 years, and smoking exposure from 30 to 20 pack-years^9^. However, lung cancer screening is still not a routine service in the UK currently. The UK National Screening Committee will meet in June 2022 to discuss this issue further.

NHS England launched a new service, the Targeted Lung Health Check (TLHC), for ever-smokers aged 55-74 years registered with a GP in autumn 2019^10^. Although this programme is expanding in geographical regions in England, patients outside the participating Clinical Commissioning Groups (CCGs) cannot access this service, which could be a potential issue of health inequality. The TLHC programme uses the Liverpool Lung Project (LLP, version 2)^11^ and the Prostate, Lung, Colorectal and Ovarian (PLCO_M2012_)^12^ cancer screening models to calculate individual patients’ risk scores as part of the eligibility criteria. Patients with risk scores higher than the thresholds (≥2.5% in LLP_v2_ or ≥1.51% in PLCO_M2012_) are considered at high risk of developing lung cancer and eligible for the TLHC programme. However, O’Dowd et al^13^ externally validated these two models using English primary care datasets (Clinical Practice Research Datalink, CPRD) and concluded that the two models did not have satisfactory discrimination (C statistic ≤0.7) and calibration. This raises the question of whether it is suitable to use these two prediction tools to select **eligible participants from the English primary care population** for TLHC and lung cancer screening. Even though the LLP_v2_ and PLCO_M2012_ models have been validated, they were developed on highly selected study samples. The LLP model was developed based on patients in Northwest England, where the age-standardised incidence of lung cancer is much higher than other regions of England^14^. The PLCO_M2012_ model was developed for the US population. The demographic and clinical characteristics of these study samples may be different from the English primary care population. Therefore, these two models may not be directly applicable to the English primary care population to select eligible participants for lung cancer screening (external validity and limited generalisability). Our team has previously developed and validated the QCancer (10-year risk) algorithms for various cancer sites^15^ using electronic health records (EHRs) from the QResearch database. A web calculator is available for public use to calculate an individual’s risk of various cancer for up to 10 years, at https://www.qcancer.org/10yr/male/ (for men) and https://www.qcancer.org/10yr/female/ (for women).

In order to provide timely research evidence for the UK National Screening Committee to expedite the decision making for population-based lung cancer screening programmes in the UK, we conducted this study to develop and validate multivariable prognostic models for lung cancer in men and women and compare the model performance of the QCancer2 (10-year risk) lung models against the other models for lung cancer screening, including the LLP_v2_ and PLCO_M2012_ models currently used in the TLHC programme in England.

## Methods

There were two stages in this study. We first updated the QCancer (10-year risk) lung model with more recent data, a larger sample size, and new predictors and validated the model. The second stage was evaluating the model performance of relevant models for lung cancer screening using the same validation dataset with the same criteria and in two patient subgroups (details below). We have published a comprehensive research protocol and statistical analysis plan for the DART-QResearch project^16^ before conducting the analyses.

### Study design, study population, and data source

This is a population-based cohort study. We used EHRs from the QResearch® database (version 45) as the data source. The QResearch® database is one of the largest healthcare databases in England, and a trusted research environment accredited by Health Data Research UK. The inclusion and exclusion criteria were similar to those in the previous QCancer (10-year risk) algorithms^15^, which allowed us to compare the model performance between the original and the updated versions of models. We included adult patients aged 25-84 years registered with general practices contributing to the QResearch database between 01 January 2005 and 31 March 2020 and excluded patients with a diagnosis of lung cancer before cohort entry. The broad age range covers the majority of the adult primary care population. It also provides great flexibility to select patients in different age groups to evaluate the model performance (Stage 2) or to perform subgroup analysis. The included patients needed to have been registered in the general practices for at least 12 months, and the general practices needed to have contributed to the QResearch database for a minimum of 12 months before the cohort entry date. This was to ensure complete data before cohort entry.

### Primary study outcome

The primary study outcome was an incident diagnosis of lung cancer recorded on one or more of the four linked data sources – primary and secondary (hospital episode statistics, HES) care databases, cancer registry (from Public Health England, PHE) and death registry (from the Office for National Statistics, ONS). We used the earliest date on any of the four records as the date of lung cancer diagnosis. We used Read/SNOMED-CT codes to identify events from the GP record, and ICD-10 codes to identify events from HES, cancer and death registries.

### Updating the QCancer (10-year risk) lung model

The predictors for the original models include age, sex, ethnicity, Townsend scores (a proxy for socioeconomic status), smoking status and intensity, alcohol status, BMI, COPD, asthma, asbestos, prior cancer, and family history of lung cancer. Two additional predictors (pneumonia^11^ and venous thromboembolism^17^) were included in the updated model. We used well-established methodologies for developing and validating the risk prediction algorithms^15 18^. Three-quarters of general practices were randomly selected for the derivation dataset and the remaining quarter for the validation dataset. Multiple imputation with chained equations was used to replace missing values for ethnicity, body mass index (BMI), alcohol and smoking status, with five imputations. We assumed that the absence of clinical information in the database indicated that the patients did not have specific health conditions, personal or family histories. Therefore, we did not perform multiple imputation for the clinical predictors. We used the imputed values in our main analyses and Rubin’s rules to combine the results across the imputed datasets^19^.

Fractional polynomials^20^ were used to model non-linear relationships between the continuous variables (age, BMI, Townsend scores) and the outcome. Cox proportional hazards models were used to estimate the coefficients for each risk factor for men and women separately, using robust variance estimates to allow for patients clustering within general practices. We initially fitted a full model with all variables for men and women, and retained the variables with a hazard ratio (HR) of <0.91 or >1.10 (for non-continuous variables) for clinical significance and statistical significance at the 0.01 level.

### How the QCancer2 (10-year risk) lung model works

We used the coefficients for each predictor from the final Cox regression as weights and combined them with the baseline survivor function evaluated for up to 10 years to derive the risk equations^15^. The baseline survivor function was estimated based on the zero values of the centred continuous variables and all binary predictors. The risk equations allow us to derive absolute risk estimates for each year of follow-up, but we had a specific focus on 5-, 6-, and 10-year risk estimates, as we intended to compare the QCancer2 (10-year risk) lung model against other models for lung cancer screening in Stage 2 of this study.

### Other prediction models for lung cancer screening to be compared against the QCancer2 (10-year risk) lung model

Toumazis et al conducted a systematic review of risk prediction models for lung cancer screening^21^. There were also empirical studies comparing some mainstream prediction models published in recent years^22 23^. We referred to these studies and included models with predictive horizons ≥5 years, as the purpose of screening is to early detect cancer from asymptomatic populations. The sojourn time for lung cancer progressing from preclinical stage (detectable by screening tests) to clinical stages is 3-6 years, longer in women than in men^24^. Detecting lung cancer at the earliest possible stage is thought to be most effective in reducing mortality, provided over-diagnosis is avoided. Therefore, prediction models for a longer period are more suitable for screening than those designed for a shorter period (e.g. 1 or 2 years) for diagnostic purposes.

We included seven models to be compared against our models, including LLP_v2_, LLP_v3_, LCRAT, PLCO_M2012_, PLCO_M2014_, Pittsburgh, and Bach. The LLP_v2_ and PLCO_M2012_ are used in the TLHC programme to select eligible participants for lung health check. LLP was updated and the newest version (LLP_v3_) was published in 2021^14^. The authors recalibrated the intercepts for each age group by sex, while the predictors and their coefficients remained unchanged between LLP_v2_ and LLP_v3_^14^. PLCO_M2014_ was updated based on PLCO_M2012_ and could be used to calculate the risk for never smokers^25^, while PLCO_M2012_ was developed for ever-smokers only. The 5-year Lung Cancer Risk Assessment Tool (LCRAT)^26^ was selected because an empirical study concluded that this model performed the best in classifying future lung cancer cases using data from three UK cohorts^23^. The 6-year Pittsburgh Predictor^27^ was included for its simplicity – only 4 predictors in the model (age and three smoking variables), with reasonably predictive accuracy. The PLCO, LCRAT, and Pittsburgh models used either all or part of the PLCO/NLST data in their models. Hence, some study samples overlapped in these models. The Bach model^28^ can predict 10-year risk, which is the same predictive horizon as our models. Most models were developed based on the US population, except for LLP using regional data from Northwest England. A total of nine prediction models were included in this study. The general information and predictors for each model are summarised in Table 1.

**Table 1.**
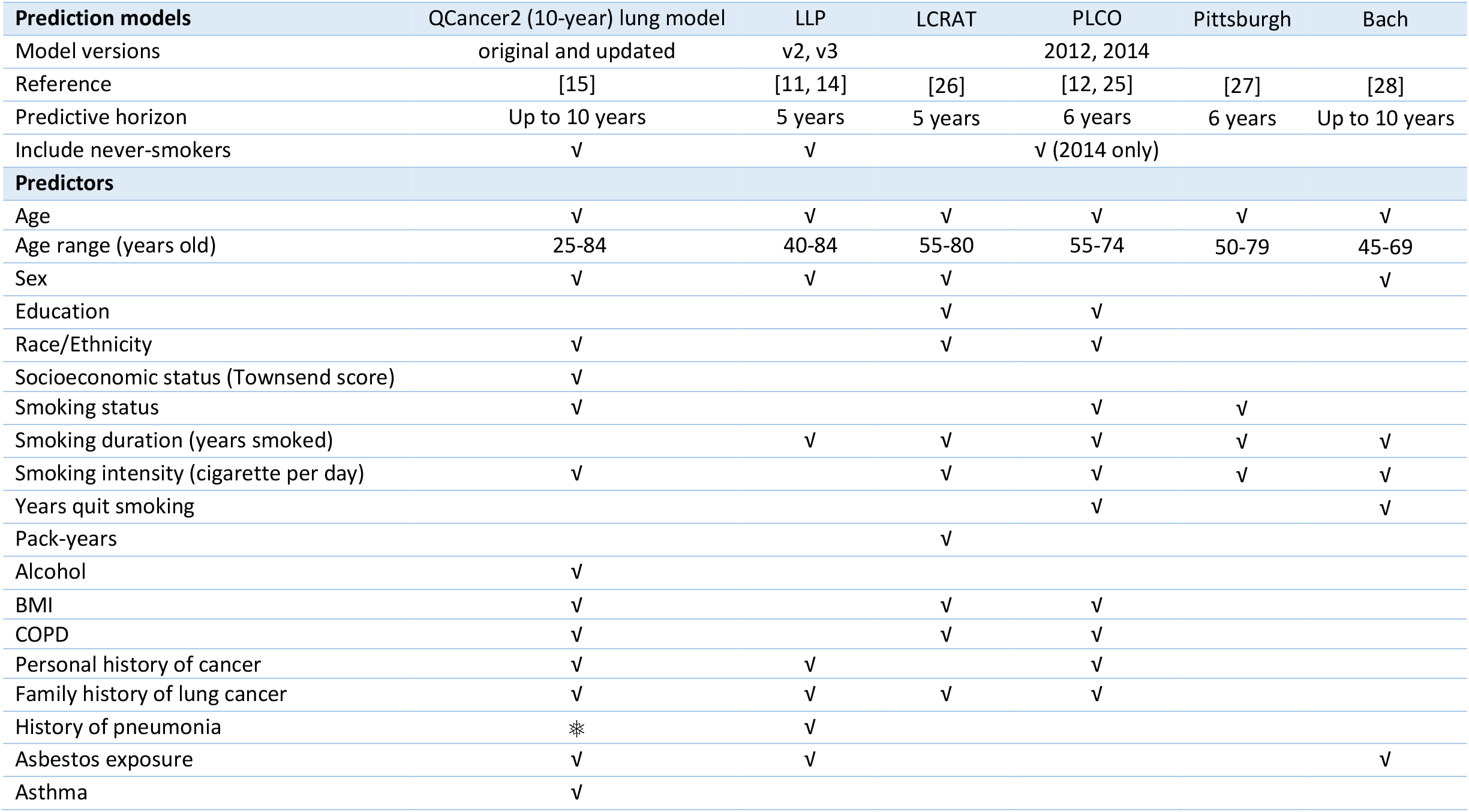
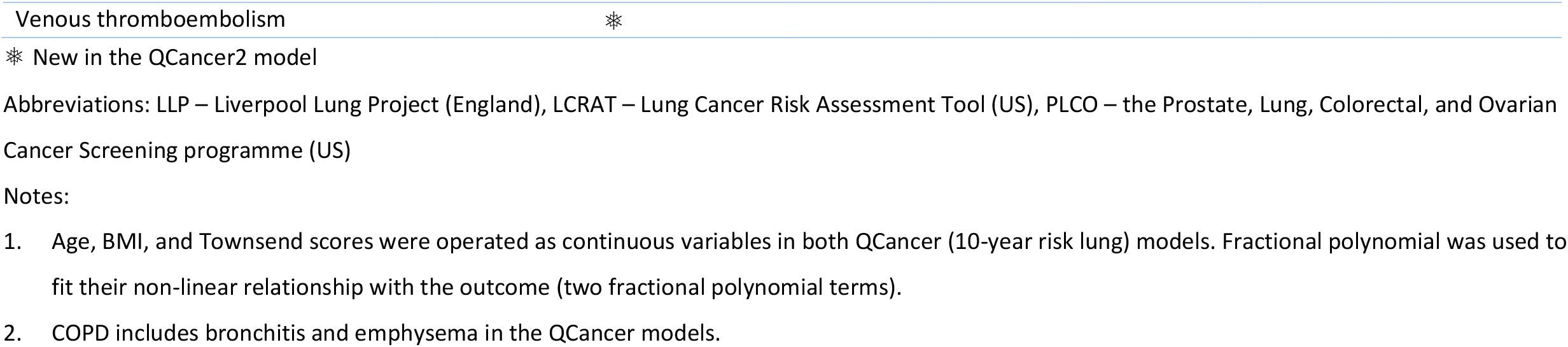
Summary of predictors in the QCancer2 (10-year risk) lung model and other prediction models

| Prediction models | QCancer2 (10-year) lung model | LLP | LCRAT | PLCO | Pittsburgh | Bach |
| --- | --- | --- | --- | --- | --- | --- |
| Model versions | original and updated | v2, v3 |  | 2012, 2014 |  |  |
| Reference | [15] | [11, 14] | [26] | [12, 25] | [27] | [28] |
| Predictive horizon | Up to 10 years | 5 years | 5 years | 6 years | 6 years | Up to 10 years |
| Include never-smokers | √ | √ |  | √ (2014 only) |  |  |
| **Predictors** |  |  |  |  |  |  |
| Age | √ | √ | √ | √ | √ | √ |
| Age range (years old) | 25-84 | 40-84 | 55-80 | 55-74 | 50-79 | 45-69 |
| Sex | √ | √ | √ |  |  | √ |
| Education |  |  | √ | √ |  |  |
| Race/Ethnicity | √ |  | √ | √ |  |  |
| Socioeconomic status (Townsend score) | √ |  |  |  |  |  |
| Smoking status | √ |  |  | √ | √ |  |
| Smoking duration (years smoked) |  | √ | √ | √ | √ | √ |
| Smoking intensity (cigarette per day) | √ |  | √ | √ | √ | √ |
| Years quit smoking |  |  |  | √ |  | √ |
| Pack-years |  |  | √ |  |  |  |
| Alcohol | √ |  |  |  |  |  |
| BMI | √ |  | √ | √ |  |  |
| COPD | √ |  | √ | √ |  |  |
| Personal history of cancer | √ | √ |  | √ |  |  |
| Family history of lung cancer | √ | √ | √ | √ |  |  |
| History of pneumonia | ✱ | √ |  |  |  |  |
| Asbestos exposure | √ | √ |  |  |  | √ |
| Asthma | √ |  |  |  |  |  |

|  |  |
| --- | --- |
| Venous thromboembolism | ✱ |

✱ New in the QCancer2 model
Abbreviations: LLP – Liverpool Lung Project (England), LCRAT – Lung Cancer Risk Assessment Tool (US), PLCO – the Prostate, Lung, Colorectal, and Ovarian Cancer Screening programme (US)
1. Age, BMI, and Townsend scores were operated as continuous variables in both QCancer (10-year risk lung) models. Fractional polynomial was used to fit their non-linear relationship with the outcome (two fractional polynomial terms). 2. COPD includes bronchitis and emphysema in the QCancer models.

### Handling EHR data for the variables in other prediction models

Due to different study designs (RCT, case-control study, or survey) and different study samples, variables in other prediction models are not necessarily available in EHR. O’Dowd et al. externally validated the LLP_v2_ and PLCO_M2012_ models using CPRD in the English population^13^. We handled most variables in a similar way as reported in their paper, except for ethnicity, the categories of smoking intensity, and the age of starting smoking. Information on patients’ ethnicity is available in the QResearch database. Therefore, we need not assume all patients were white as O’Dowd did. There are five smoking categories in the QResearch database, non-smoker, ex-smoker, light smoker (1-9 cigarettes/day), moderate smoker (10-19 cigarettes/day), and heavy smoker (≥20 cigarettes/day). We used the median number of cigarettes smoked per day for each category, i.e. 5 for light smokers, 15 for moderate smokers, and 30 for heavy smokers for the PLCO and other models, while O’Dowd et al used the lower bound of the number for moderate and heavy smoking, which may underestimate the cumulative lifetime smoking exposure. We assumed people started smoking at 15 years, as people aged ≥50 years at cohort entry (2005) were born in the 1930s-1950s. People at that time started to smoke at a much younger age and the smoking prevalence was much higher than nowadays^29^. The current legal age of smoking at 18 was set in 2007. We wrote Stata codes for the QCancer (10-year risk), LLP_v3_, and PLCO_M2014_ models, while the risk scores for the LLP_v2_, LCRAT, PLCO_M2012_, Pittsburgh, and Bach models were calculated using the “lcmodels” R package.

### Evaluation of model performance

We calculated the absolute predicted risks of individual patients by each algorithm in the validation datasets. We calculated the following measures for discrimination for each algorithm, including the 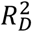(explained variation, where higher values indicate a greater proportion of variation in time to event is explained by the model)^30^, the D statistic (where higher values indicate better discrimination)^31^, and Harrell’s C statistic^32^ (similar to the area under the receiver operating characteristic curve but taking account of the censored nature of cohort data) at the prediction horizon of each algorithm (5, 6 or 10 years). These statistics were combined across the imputed datasets using Rubin’s rules. To assess calibration, we used the “pmcalplot” package in Stata^33^ to compare the mean predicted risks (x-axis) against the observed risk (obtained using the Kaplan-Meier estimates, y-axis) in twentieths by sex for each algorithm.

The three discrimination measures were calculated and calibration plots were made for the updated QCancer2 (10-year risk) lung model for the whole validation cohort (men and women aged 25-84 years). In addition, we evaluated the discrimination, calibration, and net benefits of the prediction models in two subgroups – people aged 40-84 years (including non-smokers) as subgroup 1, and ever-smokers aged 55-74 years (the criteria for the TLHC programme)^10^ as subgroup 2. There were two main reasons to set these two subgroups. Firstly, the included algorithms were developed with different inclusion and exclusion criteria (such as age range and smoking exposure), these two subgroups broadly cover the inclusion and exclusion criteria for the included algorithms so that they could be compared fairly. Secondly, we aim to provide research evidence regarding which algorithm will be the best to select eligible people for the TLHC programme if rolling out across England (through research evidence from subgroup 2). Decision curve analysis^34^ was used to evaluate the net benefit of the prediction models (clinical usefulness) for models in three predictive horizons by sex. We calculated the net benefits across a range of threshold probabilities. In general, the model with the highest net benefit at any given risk threshold is considered to have the most clinical value^35 36^.

All the analyses were conducted in the QResearch server using Stata 17.0 and R 4.1.0. We used the TRIPOD (Transparent Reporting of a multivariable prediction model for Individual Prognosis Or Diagnosis) statement to guide the conduct and reporting of this study^37 38^. A TRIPOD checklist for this article is in supplementary table S1.

### Ethical approval

This project was approved by the QResearch Scientific Committee on 8 March 2021. QResearch is a research ethics approved database, confirmed by the East Midlands – Derby Research Ethics Committee (Research ethics reference: 18/EM/0400, project reference: OX37 DART).

### Patient and public involvement

Two lay representatives from the Roy Castle Lung Cancer Foundation were involved in the project set-up phase. They reviewed our lay summary and provided feedback as part of ethical approval for this project. The lay members were not involved in the research process, but we will engage patients and stakeholders in creating accessible materials in lay language for the public to understand prediction models and risk scores of lung cancer and the benefits and harms of the TLHC or lung cancer screening when we disseminate our study findings.

## Results

### Population characteristics

Table 2 shows the baseline demographic and clinical characteristics of the study population and the incident lung cancer cases in the derivation and the validation cohorts for the updated QCancer2 (10-year risk) lung model. There were 12,991,042 patients aged 25-84 years in the derivation cohort and 73,380 people developed incident lung cancer during the follow-up period. There were 4,137,199 patients in the validation cohort and 22,838 developed incident lung cancer. The ratio of men and women was almost 1:1 in the derivation and validation cohorts, but men had a higher proportion (56%) than women (44%) in the incident lung cancer cases. The mean age for incident lung cancer cases (66 years) was 21 years older than that of the primary care population (45 years) in this study. The Townsend quintile was equally distributed in general in the derivation and validation cohorts, but had a smaller proportion in the two bottom Townsend quintiles (more deprived) in incident lung cancer cases. The percentages of ethnicity, BMI, alcohol, and smoking status recorded in the EHRs in the whole cohort were around 72%, 83%, 82%, and 93%, respectively. The proportions of comorbidities, family history of lung cancer, and any prior cancer at baseline in incident lung cancer cases were much higher than those in the primary care population.

**Table 2.** Baseline demographic and clinical characteristics of the derivation and validation cohort for the updated QCancer2 (10-year risk) model

|  | All population |  | Incident lung cancer cases |  |
| --- | --- | --- | --- | --- |
|  | Derivation | Validation | Derivation | Validation |
| Sample size (N) | 12991042 | 4137199 | 73380 | 22838 |
| Male sex, n (%) | 6476207 (49.9) | 2059175 (49.8) | 41003 (55.9) | 12768 (55.9) |
| Age, mean (SD) | 45.0 (15.6) | 45.2 (15.6) | 65.8 (10.4) | 66.0 (10.4) |
| Age groups |  |  |  |  |
| 25-29 years | 2307317 (17.8) | 718752 (17.4) | 122 (0.2) | 24 (0.1) |
| 30-34 years | 2002174 (15.4) | 628639 (15.2) | 261 (0.4) | 80 (0.4) |
| 35-39 years | 1618962 (12.5) | 517697 (12.5) | 652 (0.9) | 189 (0.8) |
| 40-44 years | 1358535 (10.5) | 436425 (10.5) | 1421 (1.9) | 479 (2.1) |
| 45-49 years | 1159196 (8.9) | 369609 (8.9) | 2908 (4.0) | 895 (3.9) |
| 50-54 years | 996141 (7.7) | 318840 (7.7) | 5212 (7.1) | 1562 (6.8) |
| 55-59 years | 913302 (7.0) | 294035 (7.1) | 8956 (12.2) | 2681 (11.7) |
| 60-64 years | 753825 (5.8) | 242618 (5.9) | 11276 (15.4) | 3466 (15.2) |
| 65-69 years | 637454 (4.9) | 204891 (5.0) | 12966 (17.7) | 4101 (18.0) |
| 70-74 years | 524814 (4.0) | 169382 (4.1) | 12710 (17.3) | 3959 (17.3) |
| 75-79 years | 431758 (3.3) | 140891 (3.4) | 10927 (14.9) | 3452 (15.1) |
| 80-84 years | 287564 (2.2) | 95420 (2.3) | 5969 (8.1) | 1950 (8.5) |
| Townsend score, mean (SD) | 0.6 (3.2) | 0.4 (3.2) | 0.4 (3.1) | 0.3 (3.1) |
| Townsend quintile, n (%) |  |  |  |  |
| Most affluent quintile (Q1) | 2830215 (21.8) | 928894 (22.5) | 16164 (22.0) | 5128 (22.5) |
| Quintile 2 | 2651740 (20.4) | 869971 (21.0) | 16110 (22.0) | 4939 (21.6) |
| Quintile 3 | 2513467 (19.3) | 823887 (19.9) | 15511 (21.1) | 4976 (21.8) |
| Quintile 4 | 2447443 (18.8) | 777626 (18.8) | 14042 (19.1) | 4212 (18.4) |
| Most deprived quintile (Q5) | 2548177 (19.6) | 736821 (17.8) | 11553 (15.7) | 3583 (15.7) |
| Ethnicity recorded, n (%) | 9381066 (72.2) | 2963779 (71.6) | 51054 (69.6) | 16200 (70.9) |
| White | 7481059 (57.6) | 2365041 (57.2) | 48418 (66.0) | 15405 (67.5) |
| Indian | 337885 (2.6) | 121854 (2.9) | 359 (0.5) | 121 (0.5) |
| Pakistani | 198742 (1.5) | 61207 (1.5) | 273 (0.4) | 69 (0.3) |
| Bangladeshi | 138942 (1.1) | 35258 (0.9) | 344 (0.5) | 72 (0.3) |
| Other Asian | 220406 (1.7) | 74806 (1.8) | 227 (0.3) | 84 (0.4) |
| Caribbean | 138224 (1.1) | 42853 (1.0) | 559 (0.8) | 183 (0.8) |
| Black African | 323946 (2.5) | 99629 (2.4) | 226 (0.3) | 59 (0.3) |
| Chinese | 104082 (0.8) | 27714 (0.7) | 108 (0.1) | 47 (0.2) |
| Other | 437780 (3.4) | 135417 (3.3) | 540 (0.7) | 160 (0.7) |
| BMI recorded, n (%) | 10797197 (83.1) | 3459976 (83.6) | 66190 (90.2) | 20841 (91.3) |
| BMI, mean (SD) | 26.5 (5.3) | 26.4 (5.2) | 26.4 (5.1) | 26.4 (5.0) |
| Smoking status recorded, n (%) | 12119036 (93.3) | 3868125 (93.5) | 70739 (96.4) | 22171 (97.1) |
| Non-smoker | 6851842 (52.7) | 2208409 (53.4) | 14352 (19.6) | 4859 (21.3) |
| Ex-smoker | 2389705 (18.4) | 756862 (18.3) | 21909 (29.9) | 6671 (29.2) |
| Light smoker (1-9 cigarettes/day) | 2150683 (16.6) | 673258 (16.3) | 24810 (33.8) | 7688 (33.7) |
| Moderate smoker (10-19 cigarettes/day) | 465255 (3.6) | 146132 (3.5) | 5150 (7.0) | 1547 (6.8) |
| Heavy smoker ($\geq 20$ cigarettes/day) | 261551 (2.0) | 83464 (2.0) | 4518 (6.2) | 1406 (6.2) |
| Alcohol status recorded, n (%) | 10531942 (81.1) | 3438730 (83.1) | 65272 (89.0) | 20649 (90.4) |
| Non-drinker | 6591000 (50.7) | 2163345 (52.3) | 41780 (56.9) | 13126 (57.5) |
| Trivial (<1 unit/day) | 2013473 (15.5) | 658992 (15.9) | 10364 (14.1) | 3260 (14.3) |
| Light (1-2 units/day) | 996128 (7.7) | 312150 (7.5) | 5340 (7.3) | 1733 (7.6) |
| Moderate (3-6 units/day) | 801199 (6.2) | 263899 (6.4) | 6467 (8.8) | 2059 (9.0) |
| Heavy (7-9 units/day) | 63064 (0.5) | 20415 (0.5) | 805 (1.1) | 314 (1.4) |
| Very Heavy (>9 units/day) | 55131 (0.4) | 16032 (0.4) | 395 (0.5) | 118 (0.5) |
| Amount not recorded | 11947 (0.1) | 3897 (0.1) | 121 (0.2) | 39 (0.2) |
| Comorbidities, n (%) |  |  |  |  |
| Any lung disease (COPD, asthma, TB, pneumonia) | 1623152 (12.5) | 519999 (12.6) | 16937 (23.1) | 5336 (23.4) |
| COPD | 182746 (1.4) | 57823 (1.4) | 9686 (13.2) | 2995 (13.1) |
| Asthma | 1295982 (10.0) | 415159 (10.0) | 7976 (10.9) | 2493 (10.9) |
| Pneumonia | 185790 (1.4) | 58429 (1.4) | 2347 (3.2) | 735 (3.2) |
| Tuberculosis (TB) | 92141 (0.7) | 30683 (0.7) | 1163 (1.6) | 404 (1.8) |
| Bronchiectasis | 30970 (0.2) | 9998 (0.2) | 467 (0.6) | 143 (0.6) |
| Asbestos exposure or asbestosis | 12714 (0.1) | 4034 (0.1) | 377 (0.5) | 127 (0.6) |
| Hypertension | 1505956 (11.6) | 488787 (11.8) | 23293 (31.7) | 7447 (32.6) |
| Venous thromboembolism | 144720 (1.1) | 46325 (1.1) | 1943 (2.6) | 639 (2.8) |
| Any cancer recorded at baseline | 299771 (2.3) | 97506 (2.4) | 5559 (7.6) | 1862 (8.2) |
| Blood cancer | 37301 (0.3) | 11969 (0.3) | 640 (0.9) | 197 (0.9) |
| Breast cancer | 87142 (0.7) | 28764 (0.7) | 1470 (2.0) | 495 (2.2) |
| Cervical cancer | 9026 (0.1) | 2944 (0.1) | 184 (0.3) | 66 (0.3) |
| Oral cancer | 6004 (0.0) | 1951 (0.0) | 211 (0.3) | 81 (0.4) |
| Ovarian cancer | 7397 (0.1) | 2369 (0.1) | 85 (0.1) | 23 (0.1) |
| Renal cancer | 20290 (0.2) | 6391 (0.2) | 623 (0.8) | 208 (0.9) |
| Uterine cancer | 7938 (0.1) | 2571 (0.1) | 119 (0.2) | 46 (0.2) |
| Family history of lung cancer | 83557 (0.6) | 28929 (0.7) | 730 (1.0) | 286 (1.3) |
| Family history of any cancer | 1436332 (11.1) | 460819 (11.1) | 6368 (8.7) | 2178 (9.5) |

### Comparison of the predictors between the original and the updated QCancer (10-year risk) lung models

The hazard ratios (HRs) for the predictors in the original and updated QCancer (10-year risk) lung models are in Table 3. The coefficients for the predictors in the updated models by sex did not change much, compared with those in the original models. Two new predictors in the updated model, pneumonia and venous thromboembolism, were both significant and had higher HRs in women than in men. Other significant predictors included ethnicity, smoking status and intensity, family history of lung cancer, COPD, asthma, prior oral, blood, and renal cancers. Compared with white, other ethnicities had a smaller HR (HR<1), except that Bangladeshi and Chinese women had an HR>1. The HR increased with the current smoking status and smoking intensity (cigarette per day) and was generally in a dose-response relationship in both sexes. The HR for moderate smokers (10-19 cigarettes/day) was smaller than that in light smokers (1-9 cigarettes/day) in men. Two smoking-related conditions, COPD and prior oral cancer, had an HR>2 in both sexes. Asbestos exposure, prior colorectal and gastric-oesophageal cancers, and drinking status, were all significant in men, but not significant in women. Such differences showed the importance of developing models separately by sex, as men and women may have different significant predictors. Even for the same predictors, the coefficients and the association between predictors and the outcome could be different between the sexes.

**Table 3.** Hazard ratios for the predictors in the original and updated QCancer (10-year risk) lung models

| Predictors | Men |  | Women |  |
| --- | --- | --- | --- | --- |
|  | QCancer2 (2022)<sup>†</sup> | Original (2015) | QCancer2 (2022)<sup>†</sup> | Original (2015) |
| White | Reference category |  |  |  |
| Indian | .504 (.439 to .58) | .503 (.438 to .578) | .676 (.58 to .788) | .674 (.579 to .786) |
| Pakistani | .597 (.515 to .691) | .595 (.514 to .689) | .718 (.554 to .931) | .717 (.553 to .929) |
| Bangladeshi | .856 (.747 to .98) | .853 (.746 to .977) | 1.04 (.827 to 1.32) | 1.04 (.824 to 1.31) |
| Other Asian | .564 (.464 to .685) | .563 (.463 to .684) | .751 (.586 to .962) | .749 (.585 to .959) |
| Caribbean | .685 (.627 to .749) | .684 (.626 to .747) | .664 (.583 to .757) | .663 (.582 to .755) |
| Black African | .526 (.429 to .644) | .525 (.429 to .643) | .689 (.556 to .853) | .687 (.555 to .851) |
| Chinese | .590 (.462 to .754) | .588 (.460 to .751) | 1.19 (.93 to 1.51) | 1.18 (.926 to 1.5) |
| Other ethnicities | .715 (.648 to .789) | .713 (.646 to .787) | .786 (.672 to .919) | .784 (.671 to .917) |
| Non-smoker | Reference category |  |  | 1 |
| Ex-smoker | 2.02 (1.82 to 2.24) | 2.02 (1.82 to 2.25) | 2.23 (2.03 to 2.44) | 2.23 (2.04 to 2.44) |
| Light smoker (1-9 cigarettes/day) | 6.59 (6.11 to 7.11) | 6.6 (6.12 to 7.12) | 5.89 (5.5 to 6.31) | 5.9 (5.51 to 6.32) |
| Moderate smoker (10-19 cigarettes/day) | 5.55 (4.94 to 6.23) | 5.56 (4.95 to 6.24) | 6.28 (5.66 to 6.95) | 6.29 (5.67 to 6.96) |
| Heavy smoker (≥20 cigarettes/day) | 9.25 (8.35 to 10.2) | 9.27 (8.37 to 10.3) | 9.9 (8.88 to 11) | 9.93 (8.9 to 11.1) |
| Family history of lung cancer | 1.44 (1.3 to 1.59) | 1.44 (1.3 to 1.59) | 1.21 (1.09 to 1.35) | 1.22 (1.09 to 1.35) |
| COPD | 2.38 (2.3 to 2.45) | 2.4 (2.33 to 2.48) | 2.42 (2.34 to 2.51) | 2.46 (2.37 to 2.54) |
| Asthma | .939 (.906 to .973) | .943 (.91 to .977) | 1.12 (1.08 to 1.16) | 1.13 (1.09 to 1.17) |
| Asbestos | 1.33 (1.2 to 1.48) | 1.34 (1.21 to 1.48) | NS | NS |
| Pneumonia | 1.21 (1.15 to 1.28) | N/A | 1.26 (1.18 to 1.34) | N/A |
| Venous thromboembolism | 1.10 (1.03 to 1.18) | N/A | 1.16 (1.09 to 1.24) | N/A |
| Prior oral cancer | 2.55 (2.18 to 2.98) | 2.55 (2.18 to 2.99) | 2.30 (1.76 to 3.01) | 2.31 (1.77 to 3.02) |
| Prior blood cancer | 1.68 (1.52 to 1.86) | 1.69 (1.53 to 1.88) | 1.90 (1.68 to 2.14) | 1.91 (1.7 to 2.16) |
| Prior renal cancer | 1.36 (1.24 to 1.49) | 1.36 (1.24 to 1.49) | 1.59 (1.36 to 1.86) | 1.6 (1.37 to 1.87) |
| Prior colorectal cancer | 1.44 (1.31 to 1.58) | 1.44 (1.31 to 1.58) | NS | NS |
| Prior gastric-oesophageal cancer | 1.27 (1.01 to 1.61) | 1.28 (1.01 to 1.61) | NS | NS |
| Prior breast cancer | N/A | N/A | 1.56 (1.48 to 1.65) | 1.57 (1.49 to 1.65) |
| Prior cervical cancer | N/A | N/A | 1.55 (1.34 to 1.79) | 1.55 (1.34 to 1.8) |
| Prior ovarian cancer | N/A | N/A | 1.27 (1.03 to 1.58) | 1.28 (1.03 to 1.58) |
| Prior uterine cancer | N/A | N/A | 1.30 (1.08 to 1.55) | 1.30 (1.08 to 1.56) |
| Non-drinker | Reference category |  |  |  |
| Trivial (<1 unit/day) | .941 (.912 to .97) | .94 (.912 to .97) | NS | NS |
| Light (1-2 unit/day) | .942 (.91 to .976) | .942 (.909 to .976) | NS | NS |
| Moderate (3-6 unit/day) | .999 (.968 to 1.03) | .998 (.968 to 1.03) | NS | NS |
| Heavy (7-9 unit/day) | 1.13 (1.05 to 1.22) | 1.13 (1.05 to 1.22) | NS | NS |
| Very Heavy (>9 unit/day) | 1.11 (1 to 1.24) | 1.12 (1 to 1.24) | NS | NS |

Notes:
† QCancer2 (2022) means the updated QCancer2 (10-year risk) lung model in this study, original (2015) means the original QCancer (10-year risk) model published in 2015<sup>15</sup>, where pneumonia and venous thromboembolism were not in the original model (N/A);
‡ The hazard ratios (HRs) in the above table are adjusted for the fractional polynomial terms for age, BMI, Townsend score, and the interaction between age and smoking status.
N/A means not applicable. NS means non-significant.

Supplementary Figure S1 shows the non-linear relationship (fractional polynomials) between the continuous variables (age, BMI, Townsend score) and the outcome (incident lung cancer cases) by sex. Supplementary Figure S2 shows the non-linear relationship between the interaction terms (age and smoking status) and the outcome by sex.

### Evaluating the model performance for prediction models for lung cancer screening

The prediction models being compared and evaluated are summarised in supplementary table S2. The descriptive statistics of the predicted risks for each model in the two subgroup populations in three predictive horizons by sex are in supplementary tables S3 and S4.

#### Discrimination

Results for the three discrimination measures (Harrell’s C, D statistics, and 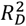) for the original and updated QCancer (10-year risk) lung models in patients aged 25-84 years are shown in Table 4. The three discrimination statistics were slightly improved in the updated model. The updated QCancer2 (10-year risk) lung model explained 65% of the variation in time to diagnosis of lung cancer. The D statistic was around 2.80 and the C statistic was around 0.9 (indicating excellent discrimination) in men and women.

**Table 4.** Discrimination statistics of the original and updated QCancer2 (10-year risk) lung models in patients aged 25-84 years by sex

| Patients aged 25-84 years | Harrell's C | D statistic | $R_D^2$ |
| --- | --- | --- | --- |
| QCancer<sup>†</sup>(2015) – Women | 0.895 (0.892 - 0.898) | 2.79 (2.75 - 2.83) | 0.650 (0.644 - 0.656) |
| QCancer (2015) – Men | 0.902 (0.900 - 0.905) | 2.77 (2.73 - 2.80) | 0.647 (0.641 - 0.652) |
| QCancer (2022) – Women | 0.897 (0.893 - 0.900) | 2.81 (2.77 - 2.85) | 0.654 (0.648 - 0.660) |
| QCancer (2022) – Men | 0.904 (0.901 - 0.906) | 2.79 (2.76 - 2.83) | 0.650 (0.645 - 0.656) |

Notes: † QCancer (2022) means the updated QCancer2 (10-year risk) lung model in this study,
QCancer (2015) means the original QCancer2 (10-year risk) lung model published in 2015<sup>15</sup>, where pneumonia and venous thromboembolism were not in the model;

The discrimination statistics for patient subgroup 1 (smokers and non-smokers aged 40-84 years) and subgroup 2 (ever-smokers aged 55-74 years) are in Tables 5 and 6, respectively, reported by sex and predictive horizons. In each patient subgroup, the QCancer2 (10-year risk) lung cancer model had the highest values for all three discrimination statistics across three predictive horizons (5/6/10 years) when compared with other prediction models. For other prediction models, in the 5-year horizon, LLP_v3_ had slightly higher Harrell’s C statistics than LLP_v2_ and LCRAT in patient subgroup 1, but comparable D statistics and 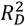 in the two patient subgroups for both sexes. In the 6-year horizon, PLCO_M2014_ had the highest Harrell’s C, but the Pittsburgh predictor had the highest D statistic and 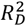in patient subgroup 1. PLCO_M2014_ consistently had the highest statistics in patient subgroup 2. In the 10-year horizon, the QCancer2 (10-year risk) lung model out-performed the Bach model. Through a comprehensive model evaluation (by sex, different age groups, and smoking exposure), we found that the three discrimination measures were often different between sexes and there was no consistent pattern of higher statistics in men or women. This again demonstrates the importance of model evaluation in different patient subgroups and sexes, rather than pooling the data together for a single summarised statistic.

**Table 5.** Discrimination statistics of models for lung cancer screening in three predictive horizons in patients aged 40-84 years (patient subgroup 1)

| Patients aged 40-84 years | Harrell's C | | D statistic | | $R_D^2$ | |
| --- | --- | --- | --- | --- | --- | --- |
| Predictive horizon: 5 year | Women | Men | Women | Men | Women | Men |
| QCancer (2022) | 0.844 (0.838 - 0.849) | 0.852 (0.848 - 0.857) | 2.243 (2.195 - 2.291) | 2.263 (2.221 - 2.305) | 0.546 (0.535 - 0.556) | 0.550 (0.541 - 0.559) |
| LLP<sub>v2</sub> | 0.647 (0.640 - 0.654) | 0.654 (0.648 - 0.661) | 1.933 (1.885 - 1.981) | 1.914 (1.872 - 1.955) | 0.471 (0.459 - 0.484) | 0.466 (0.456 - 0.477) |
| LLP<sub>v3</sub> | 0.824 (0.818 - 0.830) | 0.826 (0.821 - 0.830) | 1.992 (1.944 - 2.040) | 1.985 (1.943 - 2.027) | 0.486 (0.474 - 0.499) | 0.485 (0.474 - 0.495) |
| LCRAT | 0.595 (0.588 - 0.601) | 0.604 (0.598 - 0.610) | 1.966 (1.918 - 2.014) | 1.977 (1.935 - 2.019) | 0.480 (0.468 - 0.492) | 0.483 (0.472 - 0.493) |
| Predictive horizon: 6 year | Women | Men | Women | Men | Women | Men |
| QCancer (2022) | 0.843 (0.837 - 0.848) | 0.852 (0.848 - 0.856) | 2.241 (2.196 - 2.286) | 2.269 (2.230 - 2.309) | 0.545 (0.535 - 0.555) | 0.551 (0.543 - 0.560) |
| PLCO<sub>M2012</sub> | 0.560 (0.555 - 0.565) | 0.570 (0.565 - 0.575) | 1.456 (1.413 - 1.499) | 1.362 (1.324 - 1.400) | 0.336 (0.323 - 0.349) | 0.307 (0.295 - 0.319) |
| PLCO<sub>M2014</sub> | 0.728 (0.721 - 0.735) | 0.741 (0.735 - 0.747) | 1.426 (1.381 - 1.470) | 1.504 (1.465 - 1.543) | 0.327 (0.313 - 0.340) | 0.351 (0.339 - 0.363) |
| Pittsburgh | 0.661 (0.654 - 0.668) | 0.672 (0.666 - 0.678) | 1.954 (1.907 - 2.002) | 1.833 (1.705 - 1.960) | 0.477 (0.465 - 0.489) | 0.445 (0.411 - 0.479) |
| Predictive horizon: 10 year | Women | Men | Women | Men | Women | Men |
| QCancer (2022) | 0.842 (0.837 - 0.846) | 0.849 (0.846 - 0.853) | 2.257 (2.218 - 2.296) | 2.256 (2.222 - 2.289) | 0.549 (0.540 - 0.557) | 0.548 (0.541 - 0.556) |
| Bach | 0.691 (0.685 - 0.697) | 0.696 (0.691 - 0.701) | 1.751 (1.713 - 1.788) | 1.655 (1.622 - 1.688) | 0.422 (0.412 - 0.433) | 0.395 (0.386 - 0.405) |

**Table 6.** Discrimination statistics of models for lung cancer screening in three predictive horizons with the TLHC criteria: Ever-smoker patients aged 55-74 years (patient subgroup 2)

| Patients aged 55-74 years | Harrell's C | | D statistic | | $R_D^2$ | |
| --- | --- | --- | --- | --- | --- | --- |
| Predictive horizon: 5 year | Women | Men | Women | Men | Women | Men |
| QCancer (2022) | 0.727 (0.715 - 0.739) | 0.735 (0.726 - 0.745) | 1.925 (1.862 - 1.989) | 1.709 (1.655 - 1.762) | 0.469 (0.453 - 0.486) | 0.411 (0.396 - 0.426) |
| LLP<sub>v2</sub> | 0.647 (0.635 - 0.659) | 0.655 (0.645 - 0.665) | 1.560 (1.498 - 1.622) | 1.285 (1.232 - 1.338) | 0.367 (0.349 - 0.386) | 0.283 (0.266 - 0.299) |
| LLP<sub>v3</sub> | 0.660 (0.648 - 0.672) | 0.662 (0.652 - 0.672) | 1.634 (1.572 - 1.697) | 1.337 (1.284 - 1.390) | 0.389 (0.371 - 0.407) | 0.299 (0.282 - 0.316) |
| LCRAT | 0.642 (0.629 - 0.655) | 0.657 (0.646 - 0.667) | 1.573 (1.509 - 1.636) | 1.329 (1.275 - 1.383) | 0.371 (0.352 - 0.390) | 0.297 (0.280 - 0.313) |
| Predictive horizon: 6 year | Women | Men | Women | Men | Women | Men |
| QCancer (2022) | 0.726 (0.715 - 0.737) | 0.735 (0.726 - 0.743) | 1.910 (1.851 - 1.968) | 1.717 (1.668 - 1.767) | 0.465 (0.450 - 0.481) | 0.413 (0.399 - 0.427) |
| PLCO<sub>M2012</sub> | 0.531 (0.517 - 0.544) | 0.545 (0.534 - 0.557) | 1.281 (1.226 - 1.335) | 0.984 (0.936 - 1.032) | 0.281 (0.264 - 0.299) | 0.188 (0.173 - 0.203) |
| PLCO<sub>M2014</sub> | 0.526 (0.513 - 0.540) | 0.540 (0.529 - 0.551) | 0.734 (0.676 - 0.792) | 0.652 (0.602 - 0.701) | 0.114 (0.098 - 0.130) | 0.092 (0.079 - 0.105) |
| Pittsburgh | 0.643 (0.631 - 0.654) | 0.657 (0.648 - 0.666) | 1.577 (1.518 - 1.636) | 1.318 (1.231 - 1.406) | 0.373 (0.355 - 0.390) | 0.293 (0.266 - 0.321) |
| Predictive horizon: 10 year | Women | Men | Women | Men | Women | Men |
| QCancer (2022) | 0.724 (0.715 - 0.733) | 0.731 (0.723 - 0.738) | 1.896 (1.847 - 1.945) | 1.717 (1.675 - 1.759) | 0.462 (0.449 - 0.475) | 0.413 (0.401 - 0.425) |
| Bach | 0.575 (0.566 - 0.585) | 0.586 (0.577 - 0.595) | 1.345 (1.298 - 1.392) | 1.112 (1.071 - 1.153) | 0.302 (0.287 - 0.316) | 0.228 (0.215 - 0.241) |

#### Calibration

Calibration plots are presented in Figures 1–3. Figure 1 is the updated QCancer2 (10-year risk) lung model in three predictive horizons by sex. Figures 2 and 3 are calibration plots for the two patient subgroups. The QCancer2 (10-year risk) lung model was generally well-calibrated across different risk levels in both sexes, in three predictive horizons, in the whole validation cohort and the two subgroups. For the other prediction models, in the 5-year horizon, a common problem for the LLP_v2_, LLP_v3_, and LCRAT models in patient subgroup 2 (ever-smokers aged 55-74 years) was overestimation (the predicted risks were much greater than the observed risks) in both sexes, especially at higher risk bands. Calibration of these three models was better in patient subgroup 1, but still overestimated the risk at high-risk levels. Compared with LLP_v2_, LLP_v3_ was better calibrated in both patient subgroups. In the 6-year horizon, the PLCO_M2012_ severely under-estimated at low-risk thresholds and over-estimated at high-risk thresholds, particularly in patient subgroup 2. PLCO_M2014_ under-estimated across all risk bands in both sexes and both patient subgroups, while the Pittsburgh predictor over-estimated the risk. The Bach model was poorly calibrated in both sexes, particularly in patient subgroup 2.

**Figure 1.**
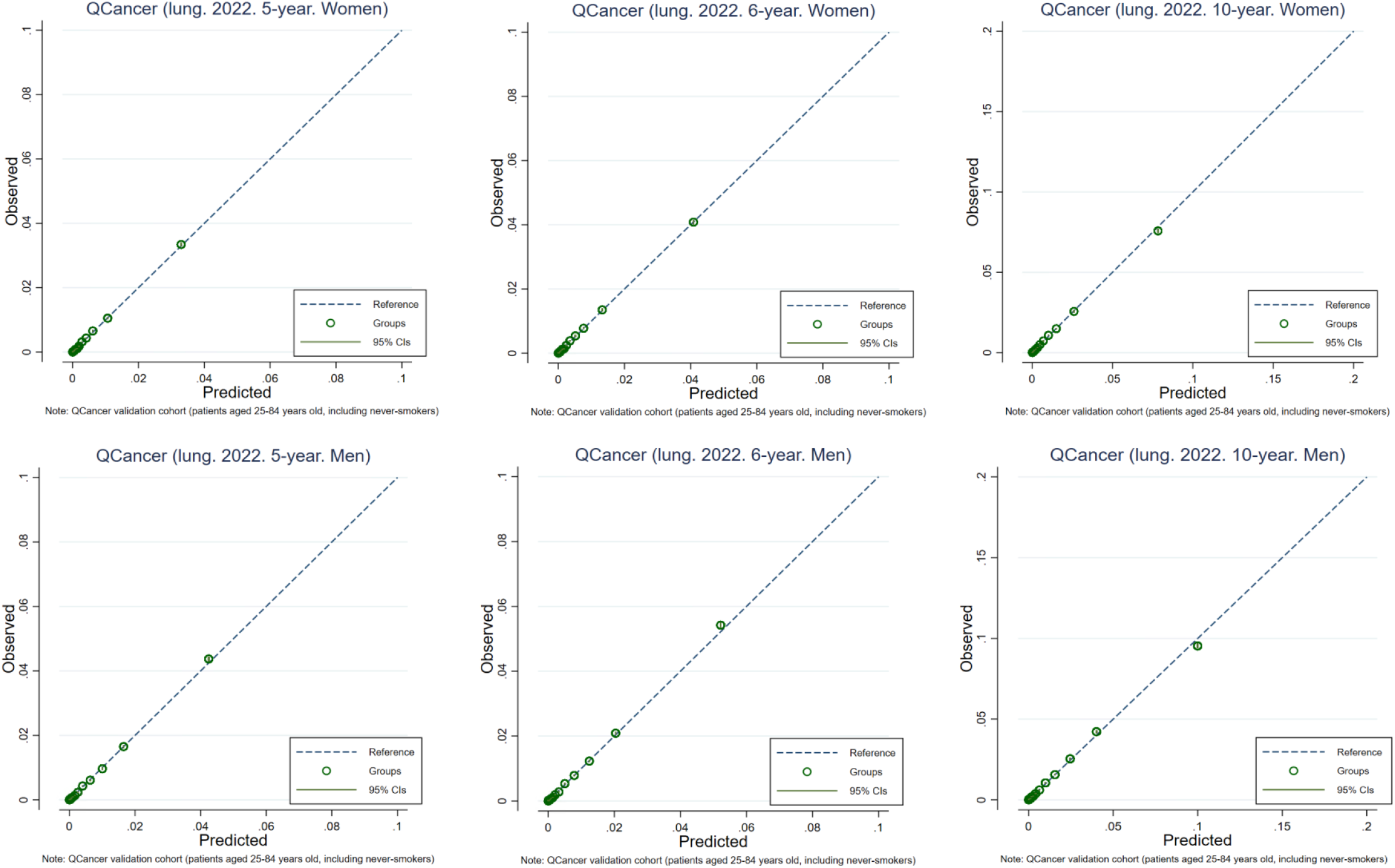
Calibration plots for the QCancer2 (10-year risk lung model) in the three predictive horizons by sex

**Figure 2.**
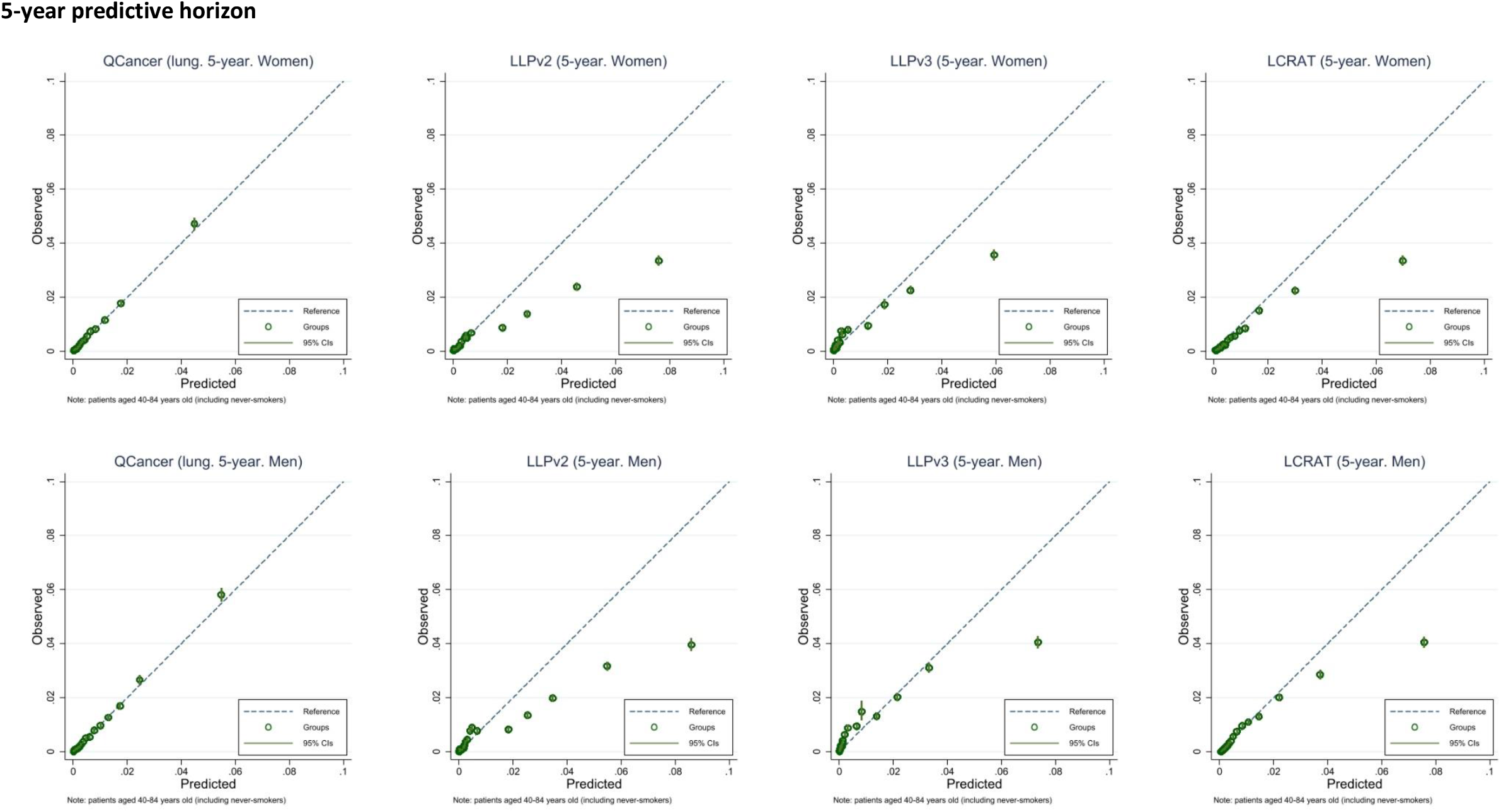
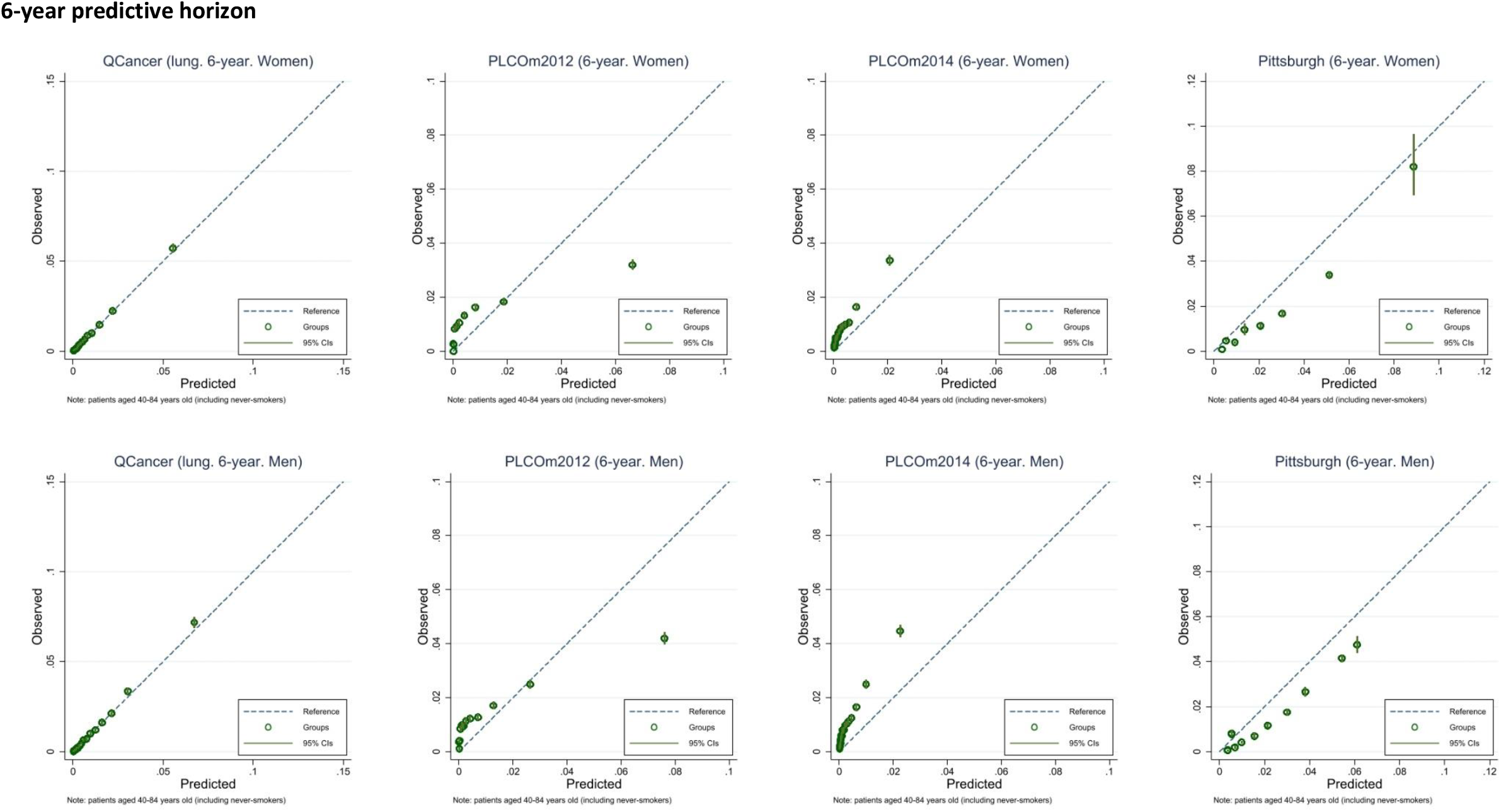
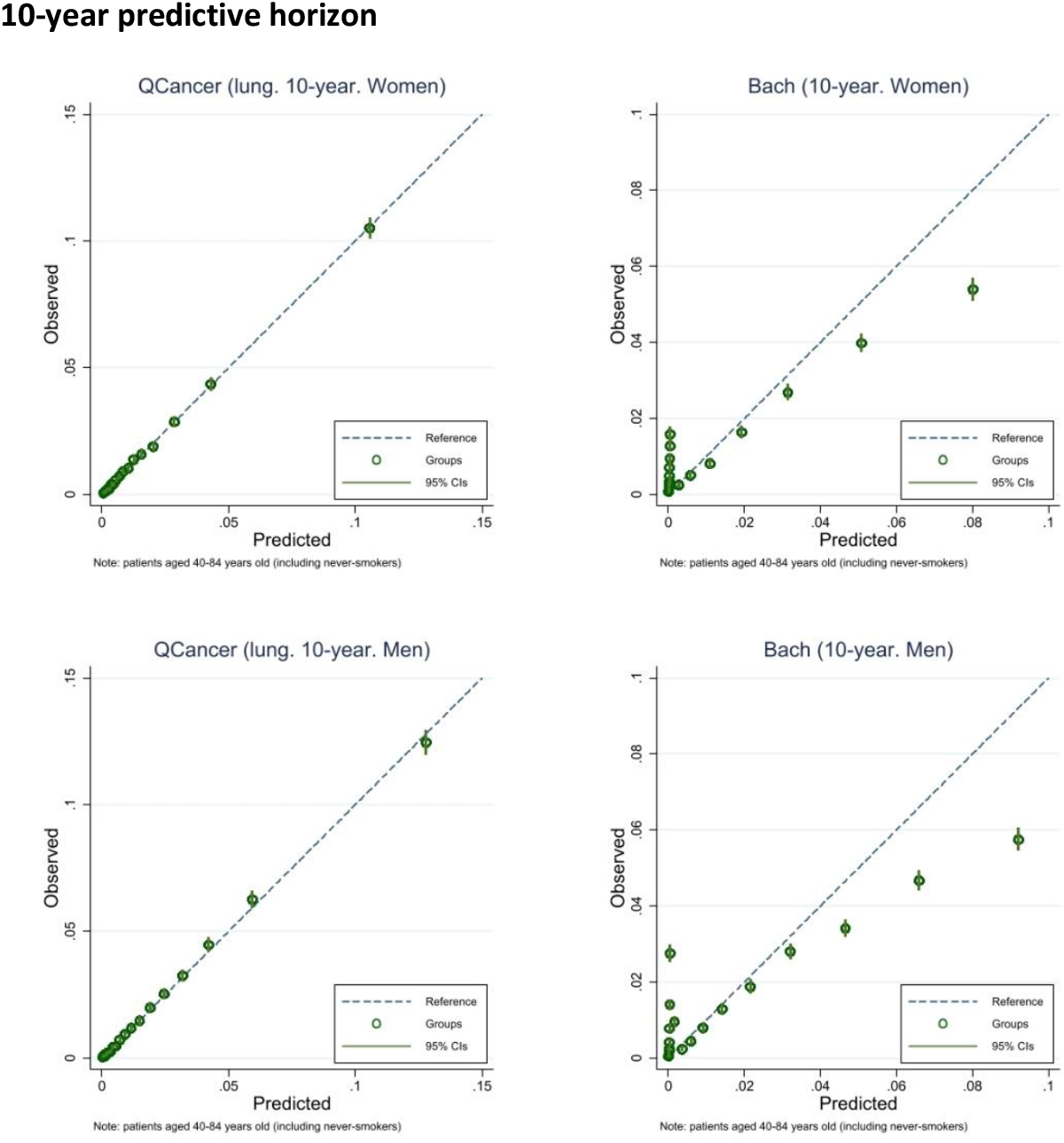
Calibration plots for smokers and non-smokers aged 40-84 years old (patient subgroup 1) in the three predictive horizons and by sex

**Figure 3.**
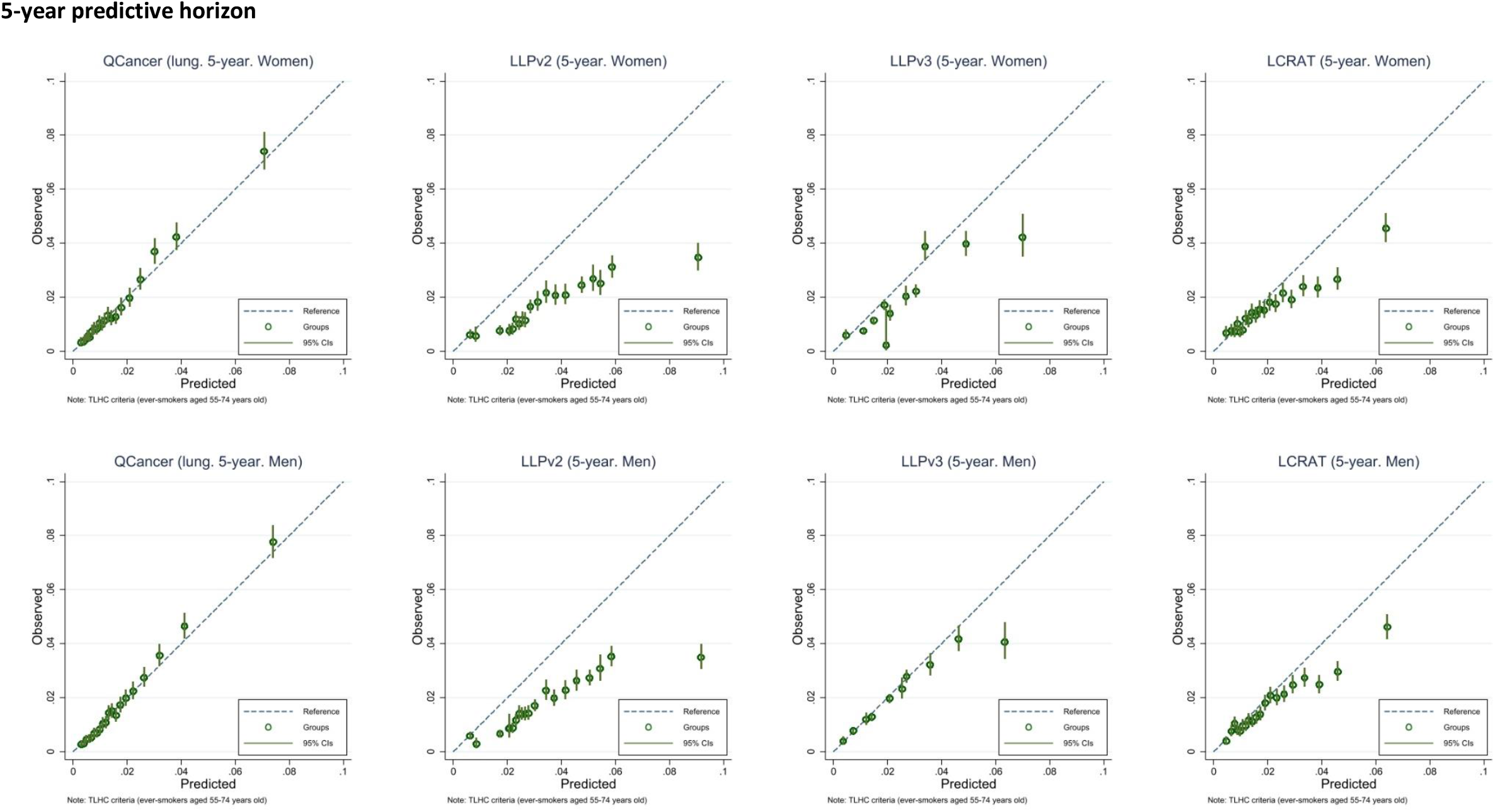
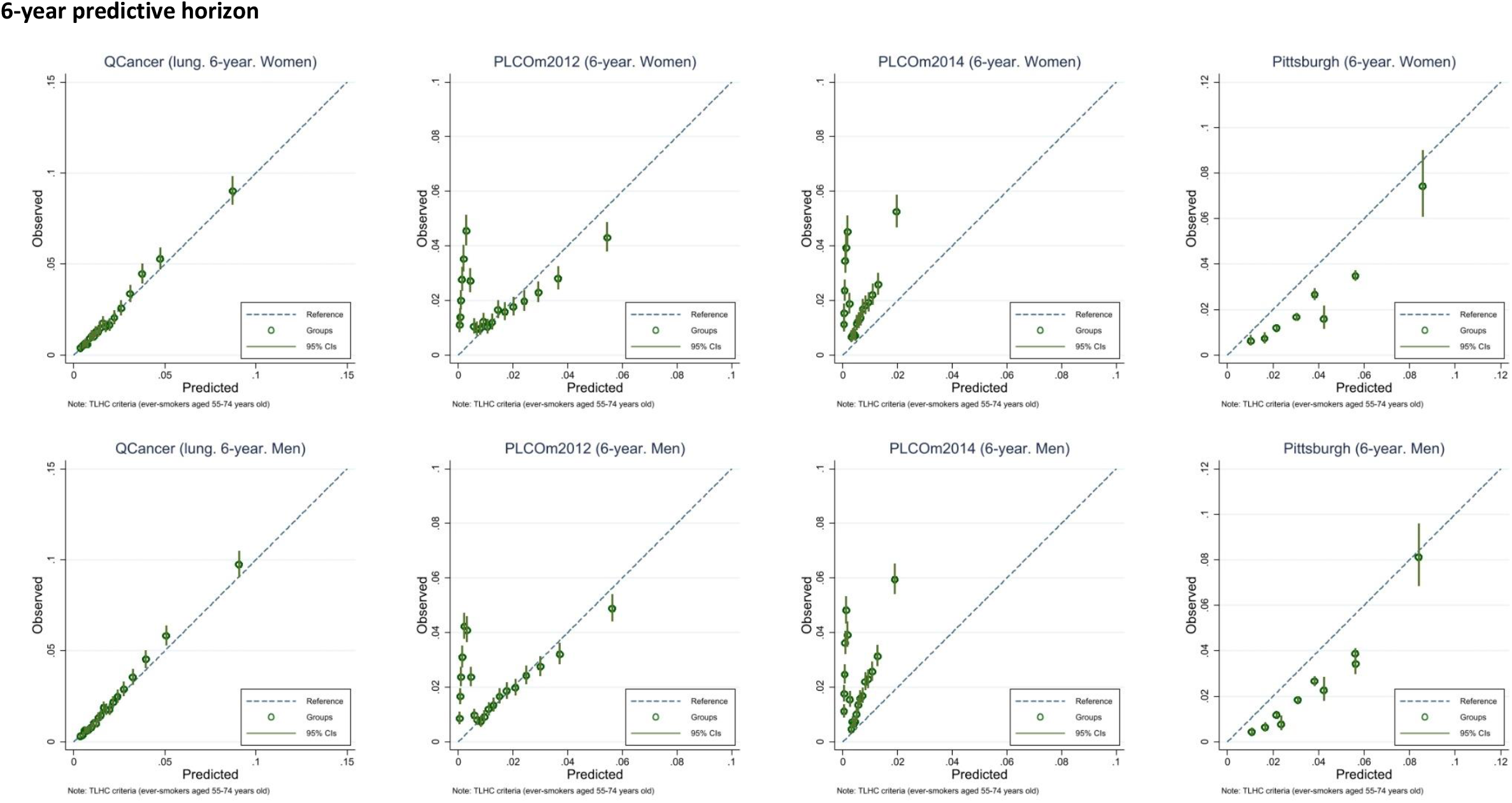
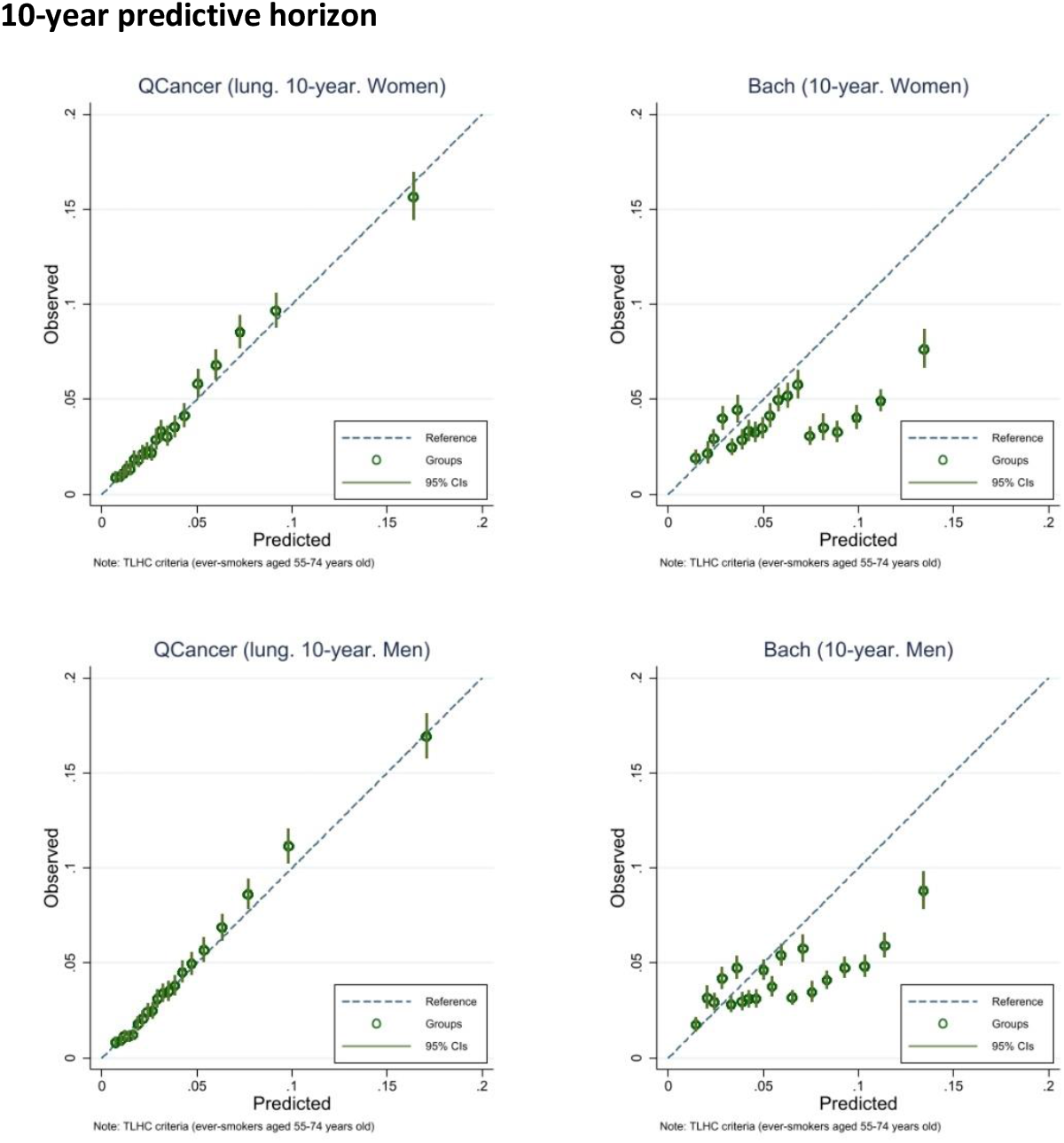
Calibration plots for ever smokers aged 55–74 years (patient subgroup 2, the targeted lung health check criteria) in the three predictive horizons and by sex

#### Decision curve analysis

Figures 4–5 show the net benefit curves for the prediction models in the two subgroups by sex in three predictive horizons. The QCancer2 (10-year risk) lung model had the highest net benefit, compared with other prediction models and strategies considering either no patients or all patients for intervention across a range of risk thresholds.

**Figure 4.**
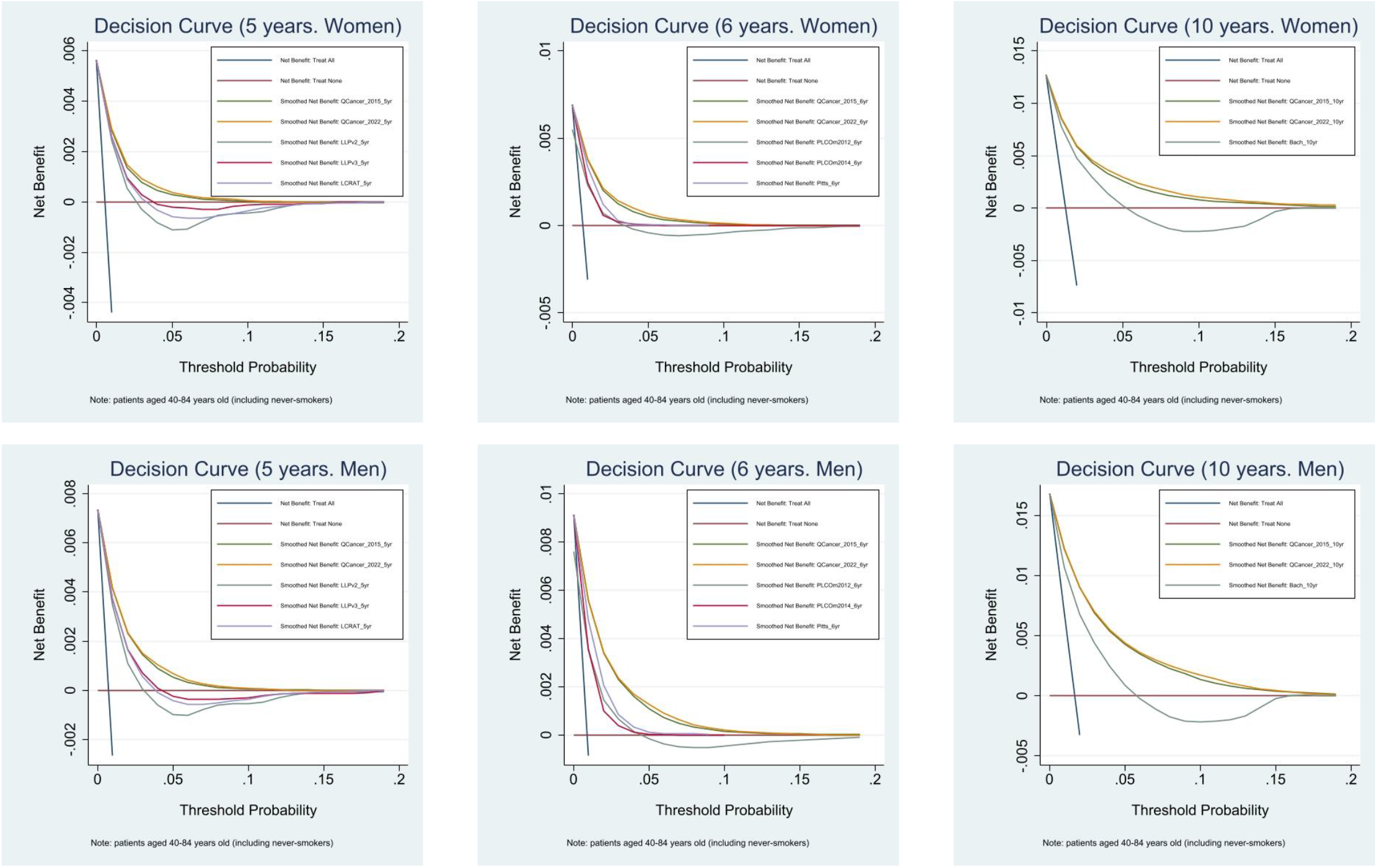
Decision curve analysis for patient subgroup 1 (smokers and non-smokers aged 40-84 years old, including never smokers)

**Figure 5.**
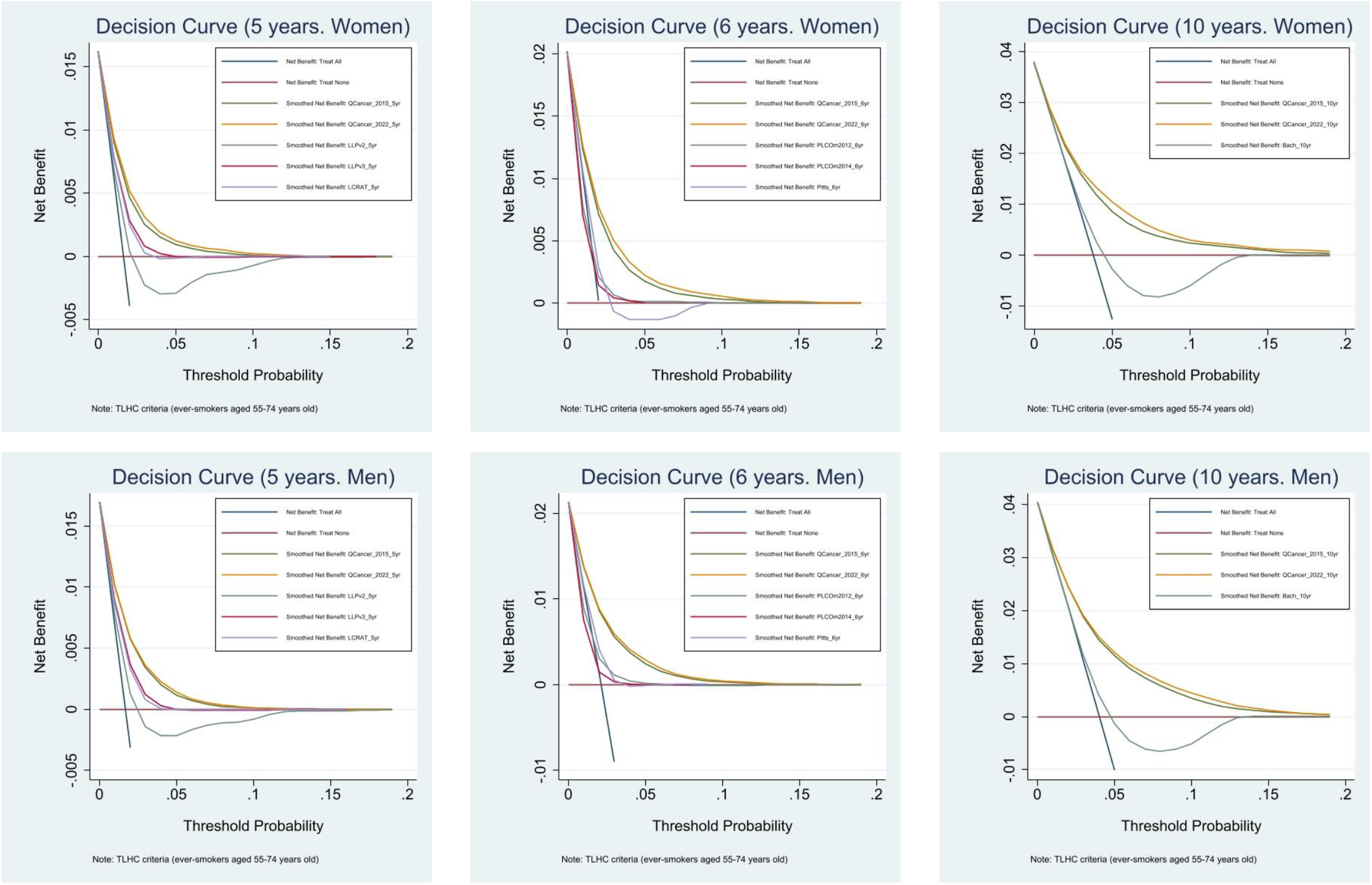
Decision curve analysis for patient subgroup 2 (ever-smokers aged 55-74 years old, the TLHC criteria)

## Discussion

### Principal findings

In this study, we developed and validated the QCancer2 (10-year risk) lung model with more recent data and a larger sample size to quantify the future risk of men and women being diagnosed with lung cancer in the next 10 years. The predictors include sociodemographic characteristics, lifestyle, comorbidities, family history and personal history of other cancers. The updated model performed slightly better than the original version. We compared the QCancer2 (10-year risk) lung model against other prediction models and found that the QCancer2 (10-year risk) lung model had the best performance in discrimination, calibration, and net benefits in two subgroups across three predictive horizons.

### Comparison with existing literature

The QCancer (10-year risk) models are ***prognostic*** models, which can estimate an individual patient’s ***future*** risk of cancer while the patient has not yet presented any symptoms. Prognostic models are more suitable for early detection of cancer through screening in asymptomatic populations, while diagnostic models (known as QCancer) aim to predict the risks of existing cancer in patients already present with symptoms that may indicate cancer.

Compared with the original model using a total of 6.6 million patients aged 25-84 years and 13,570 incident lung cancer cases from 753 practices from 1 January 1998 to 30 September 2013 in the deviation and validation cohorts, the sample size of the whole population and the incidence lung cancer cases in the updated model increased substantially, due to the expansion of the QResearch database in recent years. We have conducted a more thorough assessment of model performance in this study. ten Haaf et al. mentioned little attention has been given to sex-specific risk-stratification in current practice^39^. We developed and validated our models by sex and found that the predictors differed between men and women. Even for the same predictors, their coefficients may be different. The histological subtypes and preclinical duration of lung cancer may vary between sexes^24 39^. It may need to consider sex-specific risk thresholds and screening intervals for lung cancer screening.

When choosing risk prediction models for population-based screening programmes, we need to consider whether the models can be applied to the targeted population (external validity and generalisability), as models may be developed using a specific study sample, with limitations in study design, data, or internal validity of the study findings. Some models may be developed using highly selective study samples or populations, whose baseline risk was higher than the general population. The LLP model had limitations in study design and study sample (case-control study using regional data)^11^. The PLCO_M2012_ model was developed using data from the US population^12^. When externally validating these two models using CPRD, O’Dowd found that both models underestimated the risk in patients at low predicted risk (observed risk>predicted risk) but overestimated the risk in patients at higher risk (observed risk<predicted risk, cut-offs at around 1% for both models)^13^. The C statistics (equivalent to AUC in O’Dowd’s report) in this study were even lower. Possible explanations included study sample selection (two patient subgroups in this study vs ever-smokers aged 50-80 years), availability of information, population and geographical coverage in different EHR databases (QResearch vs CPRD), different ways of handling the variables unavailable from the EHR database (described in the methods section), sample size and study period (this study had a larger sample size and a longer study period). Despite these differences, both studies reached the same conclusion that the LLP_v2_ and PLCO_M2012_ models did not have satisfactory discrimination and were not well-calibrated when externally validated using different English primary care databases. They may not be directly applicable to the English primary care population.

Robbins et al. evaluated the model performance of lung cancer risk models (LLP_v2_, LLP_v3_, LCRAT, LCDRAT, PLCO_m2012_, and Bach) in current and former smokers aged 40-80 years using three UK cohorts (UK Biobank, EPIC-UK, and Generations Study) to define the eligibility for lung cancer screening^23^. We did not include LCDRAT in our study, as this model predicts lung cancer death. The AUC values for the models in the two patient subgroups (patients aged 40-74 years and 55-74 years, 0.73-0.82) were much higher in the three cohorts than those in our study. For calibration, all models overestimated risk in all cohorts. The ratio of expected vs observed (E:O) was between 1.20 (LLP_v3_) and 2.25 (LLP_v2_). We found both overestimation and underestimation appeared in our study samples. Robbins et al. emphasised the importance of validating prediction tools in specific countries. We agreed with this point.

### Strengths and limitations of this study

#### Strengths

Informed by the existing research evidence and clinical expertise, we included as many relevant predictors as possible when we updated the QCancer2 (10-year risk) lung model. Using contemporaneous primary care EHRs to develop and validate risk prediction models is likely to have great face validity, generalisability, and applicability to the whole primary care population, compared with study designs like RCT or survey, or data from other countries (like the US). EHRs in the UK have a high level of accuracy and completeness of sociodemographic information and clinical diagnoses. Clinical and diagnostic information recorded by healthcare professionals should be more accurate than collecting such information from patients using surveys or interviews. Therefore, it can reduce information and recall bias. In addition, our study also benefits from a large sample size and a long duration of follow-up. The population in the QResearch database is representative of the whole English primary care population, in terms of age, sex, ethnicity, socioeconomic deprivation, smoking exposure, and geographical coverage. Therefore, it minimises selection and respondent bias. The QCancer2 (10-year risk) lung model could be applied to the whole English primary care population. We would like to consider our QCancer2 (10-year risk) lung models as inclusive algorithms. They were designed for the adult primary care population with a wide age range (25-84 years) and various smoking statuses (non-, current, or ex-smokers). They are “live” and flexible risk prediction models, which can be updated based on the previous version with more recent data, as the population changes over time. Physicians and the public can use our models to calculate and understand an individual’s risk of developing lung cancer for each year of follow-up, for up to 10 years, which is more flexible than models predicting a fixed predictive horizon such as 5-year for LLP and 6-year for PLCO. The QCancer2 (10-year risk) lung models were developed and validated in men and women separately, with some different predictors in the men and women models. Therefore, the models allow sex-specific risk stratification. Compared with other models, this is a unique strength. The fact that the QCancer2 (10-year risk) lung models outperform other prediction models in the ever-smokers aged 55-74 years (subgroup 2) demonstrates that our model is robust and suitable for selecting eligible patients for lung cancer screening. This bonus point makes our models more attractive for clinical application and improving population health.

Finally, we pre-registered the research protocol and statistical analysis plan in the public domain. We used robust and advanced statistical methods and followed the recommendations of the TRIPOD guidelines when we developed and validated our models and reported the study findings.

#### Limitations

Smoking is a strong predictor of lung cancer. When primary care providers ask the smoking status and intensity, some patients may feel ashamed and under-report their smoking intensity than their actual situation. This could affect the accurate estimation of the association between smoking exposure and lung cancer. But there is little that researchers could do about this. Family history of cancer may have been recorded opportunistically rather than systematically in the EHR. Patients with a family member diagnosed with cancer may be more likely to report this to their GPs and for this to be recorded, which would cause information bias. Not all relevant information in other prediction models is available from EHRs. Therefore, we needed to make reasonable assumptions and adapt the situation for the EHR in the English population when we calculated the risk scores using other prediction models. If we could not find values from reliable sources or references, we used the median values or categories, as O’Dowd et al^13^ did in their study. The unavailable information and the way we handled the variables may inaccurately estimate the risk scores for individual patients in other prediction models, which could either overestimate or underestimate the risk. This may consequently influence the evaluation of model performance for individual models. Another limitation is that we did not use external data to validate QCancer (10-year risk) lung models. It is possible to use another data source (e.g. CPRD) and the internal-external cross-validation approach^40^ to externally validate our model in future studies, subject to funding availability.

### Implications for practice and health policy

As a universal service with a very high population coverage in the UK, primary care has great potential and opportunity for early detection and diagnosis of cancer, and GPs could be strong advocates for the TLHC programme and lung cancer screening. We have demonstrated the model performance in the QCancer2 (10-year risk) lung model. More accurate risk estimation has an impact on risk stratification at the population level and risk communication with the patients. The implementation of the QCancer2 (10-year risk) lung model in primary care computer systems could facilitate patient-GP discussion about the risks and benefits of preventive interventions for individual patients. This could improve patients’ awareness of their health status and risk level, which in turn might increase their willingness to participate in the TLHC programme or lead to behavioural changes such as considering smoking cessation.

The updated QCancer2 (10-year risk) lung model allows batch-mode processes that use existing information in EHRs at each practice to facilitate the selection of eligible patients at high risk for the TLHC programme or lung cancer screening. Compared with using questionnaires to collect information from individual patients and calculating risk scores to check their eligibility, the batch-mode process is more efficient. It can stratify the primary care population based on risk level for better target in prevention or screening programmes. It can not only substantially reduce human resources and costs, but also save time and streamline the administrative process for better patient experience and increased patient satisfaction.

We comprehensively compared nine prediction models using the same validation dataset in this study. We evaluated model performance in two patient subgroups, which broadly covered the inclusion and exclusion criteria of the study sample in each model and the eligibility criteria for the TLHC programme. The LLP_v2_ and PLCO_M2012_ may not be the best models to select eligible primary care patients for the TLHC programme, due to their unsatisfactory model performance in patient subgroup 2. Through such a thorough assessment in this study, we provide research evidence to assist policymakers in considering the age range and smoking exposure for lung cancer screening and increase equality in health services access. In the end, more patients can be diagnosed at earlier stages and through screening-detected route, which can lead to better survival outcomes and reduced lung cancer mortality in the UK. Considering the sizable incidence and mortality of lung cancer at the population level, the improvement in lung cancer could substantially contribute to the UK government’s ambition that 75% of people with cancer will be diagnosed at early stages by 2028^41^. Meanwhile, patients may benefit from primary or secondary prevention of other cardiorespiratory diseases through participating in the TLHC programme. We do not recommend a risk threshold for our models for lung cancer screening, as this needs to consider the balance between benefits and harms, cost-efficiency, health resources, accessibility and health equality, and the potential impact of the screening program at the population level. Our colleagues in another strand of our work package will conduct a separate study to evaluate the cost-effectiveness and determine the risk thresholds for lung cancer screening in England.

## Conclusion

Developed, updated, and validated using primary care EHRs, the QCancer2 (10-year risk) lung model is robust and flexible. It can estimate an individual adult patient’s risk of lung cancer diagnosis during each year of follow-up, for up to 10 years. It has the best performance among other prediction models for lung cancer screening in discrimination and calibration for both sexes across three predictive horizons (5/6/10 years). It also has the highest net benefit. The QCancer2 (10-year risk) lung model is more suitable for selecting patients at high risk from the English primary care population for lung cancer screening using LDCT than the currently used LLP_v2_ and PLCO_M2012_ models in the TLHC programme.

## Data Availability

To guarantee the confidentiality of personal and health information of patients, only the named authors (WL and JH-C) have had full access to the data during the study, in accordance with the relevant licence agreements. Information on access to the QResearch data is available on the QResearch website (www.qresearch.org).

## Declarations

WL and JH-C have full access to all data in this study and take the responsibility for the integrity of the data and the accuracy of data analysis. To guarantee the confidentiality of personal and health information of patients, only the named authors have had full access to the data during the study, in accordance with the relevant licence agreements. Information on access to the QResearch data is available on the QResearch website (www.qresearch.org).

JH-C is the guarantor for the study and affirms that the manuscript resulting from this work is an honest, accurate, and transparent account of the study being reported; that no important aspects of the study are omitted; and that any discrepancies from the study protocol and statistical analysis plan^16^ have been explained.

## Authors’ contributions and consent for publication

FG and JH-C secured the funding. FG is the chief investigator of the DART project, and JH-C is the joint package lead (WP6 – Primary care, population health, and health economics). JH-C and WL contributed to the study conceptualisation. WL specified the data and led on the ethical approval. WL designed the statistical analysis plan and drafted the whole research protocol and statistical analysis plan, with methodological input from JH-C and CC, and clinical and contextual input from JH-C, DB, FG, and JB. WL and JH-C wrote the codes and performed the analyses. CC had substantial contributions to the statistical methodology. WL drafted the whole paper. All authors contributed to the interpretation of the results and revision of the manuscript, and approved the final version of the manuscript for publication.

## Acknowledgements

We thank the two lay members from the Roy Castle Lung Cancer Foundation who reviewed our lay summary of the DART-QResearch Project for ethical approval and provided very helpful feedback. We acknowledge the contribution of the patients and the general practices who contribute to QResearch® and EMIS (Egton Medical Information Systems) Health and the Chancellor Masters & Scholars of the University of Oxford for expertise in establishing, developing and supporting the QResearch database. The Hospital Episode Statistics data used in this analysis are Copyright © (2022) to the Health and Social Care Information Centre and re-used with the permission of the Health and Social Care Information Centre and the University of Oxford. All rights reserved. This project involves data derived from patient-level information collected by the NHS, as part of the care and support of cancer patients. The data is collated, maintained and quality assured by the National Cancer Registration and Analysis Service, which is part of Public Health England (PHE). Access to the data was facilitated by the PHE Office for Data Release. The death registration data are provided by the Office for National Statistics. NHS Digital, Public Health England, and the Office of National Statistics bear no responsibility for the analysis or interpretation of the data.

The views expressed in this manuscript are those of the authors. None of the acknowledged organisations or funding bodies has been involved in any research process, including study design, data specification, statistical analysis, interpretation of results, preparing manuscripts, or decision to publish.

## Study and authors’ information

Project website: www.dartlunghealth.co.uk

## Twitter

@dartlunghealth (the DART project),

@WLiao_Ox (Weiqi Liao),

@JudithBurchardt (Judith Burchardt),

@JuliaHCox (Julia Hippisley-Cox)

